# Toward less intrusive pubertal assessment: longitudinal evaluation of tanner and non-tanner metrics in East African adolescents

**DOI:** 10.64898/2026.06.17.26355904

**Authors:** Aggrey J. Anok, Jessica L. Prodger, Jodie L. White, Daniel E. Park, Ping Yang, Simon Peter Bukenya, Stephen Kiboneka, Mathias Agaba, Mary J. Nalubowa, James Nnamutete J, Edward N. Kankaka, Nathan O. Weber, Cindy M. Liu, Rupert Kaul, Godfrey Kigozi, Aaron AR Tobian, Ronald M. Galiwango

## Abstract

**Background:** Accurate pubertal assessment is essential in pediatric endocrinology and adolescent health research. While Tanner staging remains the gold standard, its subjective nature and invasive genital examination limit feasibility and acceptability, especially in longitudinal studies and culturally sensitive settings. This study evaluated less intrusive pubertal assessment combinations that maintain discriminative accuracy.

**Methods:** We conducted a longitudinal study among 200 uncircumcised, sexually naïve males aged 15–17 years in Southwestern Uganda, with quarterly follow-up over three years. Clinicians assessed Tanner staging metrics (pubic hair, testicular volume, penile length, scrotal color), axillary hair, and serum testosterone. Markov transition models estimated Tanner stage progression. Ordinal logistic regression and area under the receiver operating characteristic curve (AUC) analyses quantified discriminative performance of individual and combined metrics.

**Results:** At baseline, participants were distributed across Tanner stages II (6.0%), III (13.5%), IV (55.0%), and V (25.5%). Among individual metrics, pubic hair distribution best predicted overall Tanner stage (AUC=0.867), while penile length was least predictive (AUC=0.833). The full four-metric Tanner model achieved high discrimination (AUC=0.993). However, a less intrusive combination of pubic hair and scrotal color achieved comparable discrimination (AUC=0.942), improving to AUC=0.953 with axillary hair and age. Markov modeling demonstrated frequent bidirectional transitions between Tanner stages IV and V, reflecting variability in longitudinal staging.

**Conclusions:** A minimally intrusive assessment combining pubic hair, scrotal color, axillary hair, and age reliably predicts pubertal stage, offering an acceptable alternative to traditional Tanner staging for research and surveillance contexts where genital manipulation is impractical or unethical.

## Introduction

Puberty is a critical stage of life during which a child undergoes physical, physiological, and hormonal changes that transition them into adulthood [1]. These changes affect metabolism, body function, and personality traits, potentially influencing long-term health outcomes [2]. This period can shape dietary habits and physical activity preferences, thereby impacting future risks for non-communicable diseases (NCDs) such as diabetes, hypertension, and depression [3]. Given the growing burden of NCDs, accurately tracking pubertal development is essential to inform appropriate clinical and non-clinical interventions for holistic adolescent care [4,5].

The Tanner Sexual Maturity Rating system, developed by James Tanner in 1962, remains the gold standard for assessing adolescent physical development [6]. This system evaluates key primary and secondary sexual characteristics in boys and girls. For boys, these include observation metrics (pubic hair distribution, changes in scrotal skin color and texture) and genital manipulation metrics (penile length, testicular volume). Each characteristic is scored on a scale from 1 to 5, culminating in a final Tanner stage classification: prepubertal (stage 1), early pubertal (stage 2), mid-pubertal (stage 3), late pubertal (stage 4), and adult/post-pubertal (stage 5). However, concerns have been raised regarding the Tanner staging system’s reliability, with false-positive rates ranging from 2% to 19% and false-negative rates from 1% to 18% [7].

Furthermore, Tanner staging includes intrusive genital manipulation to determine penile length and testicular volume, which can be particularly disturbing for young. Self-assessment of maturity staging has been proposed as an alternative and found to have some utility within specific gender and puberty stages [8,9]. However, there is limited data from Africa and some studies have indicated inaccuracies of maturity self-assessment, including possible anxiety and self-esteem deficits, thereby partly reinforcing the need for clinician-performed evaluations [10,11].

There is a pressing need for a reliable, minimally intrusive yet accurate tool to assess pubertal development for both clinical and research purposes. Exploring alternative staging systems or refining the Tanner model could enhance reliability while reducing concerns around privacy and emotional distress. In this analysis, we leverage data from a study tracking genital microbiome and immunological changes associated with reproductive maturation in a cohort of sexually naive, HIV-negative Ugandan adolescent boys. As part of this study, we examined changes in tanner and non-tanner sexual maturity markers over three years to identify potential metrics for a minimally intrusive pubertal assessment tool.

## Materials and methods

### Study design and participants

The objective of this analysis was to identify combinations of pubertal characteristics that preserve discriminative accuracy of pubertal stages while minimizing the need for genital manipulation. Analyses were performed on data collected during a longitudinal, observational study of 200 pre-sexual adolescent boys, aged 15–17, in Southwestern Uganda. Participants were identified through ongoing Rakai Community Cohort surveys [12,13] or were referred by community health educators. Enrollment occurred between September 2017 and February 2018 with study visits every three months for up to three years (maximum 13 visits including enrollment visit). Inclusion criteria required parental consent, individual assent, age between 15-17 years at enrollment, HIV-negative status, and self-reported sexual naivety. Participants with phimosis were excluded. Written, informed parental consent and participant assents were obtained for all participants.

The study was approved by the institutional review boards at the Uganda Virus Research Institute, Uganda National Council for Science and Technology, Johns Hopkins University, George Washington University and Western University.

### Study procedures

HIV counseling, testing, and completion of a baseline socio-behavioral questionnaire on sexual activity was performed at enrollment. Two study clinicians conducted physical and genital examinations at each study visit, without reference to prior visit measurements and no participant was seen by both clinicians at any given visit. General physical findings were recorded, and genital maturation was assessed using the Tanner sexual maturity rating. In the supine exposed position, penile length was measured using a stiff paper strip aligned from the base to the tip and compared against a measuring tape. Length was categorized into four levels: <3cm (Category 1), 3-6cm (Category 2), >6-10cm (Category 3), and >10-15cm (Category 4). Testicular volume was assessed using an Erler-Zimmer orchidometer (12 beads), categorized into five levels: <1.5mL (Category 1), 1.6-<6mL (Category 2), 6-<12mL (Category 3), 12-20mL (Category 4), and >20mL (Category 5). Scrotal color was staged using a Tanner chart (scale 1-3), and pubic hair distribution was graded on a scale of 1-5 [6]. Axillary hair distribution, a non-Tanner metric, was scored using Wolfsdorf staging to assess its utility in alternative sexual maturity ratings [14]. The Wolfsdorf staging tool was incorporated into the study protocol nine months after study initiation, with approval from the Institutional Review Board. The amendment was made in response to concerns that scrotal skin color may not reliably indicate physical maturation in African adolescents and that axillary hair distribution would enhance assessment of pubertal development in this population. As such, study visits that occurred prior to this amendment did not include collection of axillary hair data.

Participants also provided 12mL of peripheral venous blood, which was processed into serum. Aliquots were frozen at −80°C for later batch testing of testosterone levels using the Elecsys Testosterone II electrochemiluminescence immunoassay on the Roche Diagnostics cobas e 602 analyzer with a lower limit of detection of 2.5 ng/dL. Participants remained enrolled regardless of sexual debut. Loss to follow-up was defined as the inability to trace a participant until study completion.

### Statistical Analysis

Prior to analysis, data transformation was conducted to standardize variables and address data missingness. Individual Tanner metrics were initially scored on their original scale, then transformed for analysis. For testicular volume, the original 5-point scale was reduced to a 4-point scale by excluding the lowest category (<1.5mL), as this stage was not observed in our sample. The highest two categories (12-20mL and >20mL) were collapsed into a single category due to the rarity of participants with testicular volumes <20mL. Penile size was subsequently collapsed to a 3-point scale by combining the categories 3 and 4 categories, as penile length greater than 10-15cm (category 4) is rarely observed among boys in the sampled age range. All tanner metrics were linearly rescaled to a common 1-5 scale to ensure equal weighting. A composite Tanner stage score was calculated as the arithmetic mean of all four rescaled scores, providing a continuous measure of the tanner score ranging from 1 to 5.

Missing axillary hair values were imputed using a linear mixed-effects model with age as a fixed effect and participant ID as a random intercept. The model was fitted on complete cases and used to predict missing values, which were then rounded and scaled to the 1-5 Tanner scale range. This imputation method leveraged the expected positive association between chronological age and pubertal development while accounting for individual variation in pubertal timing through participant-specific random intercepts.

Descriptive statistics, including frequencies and medians, were used to summarize participant characteristics such as age, occupation, education, and Tanner stage. Kendall’s tau-b, a non-parametric measure of monotonic association between ordinal and continuous variables, was used to assess correlations between age and Tanner or non-Tanner metrics, including serum testosterone levels. Markov transition models estimated the probability of progression between Tanner stages, providing insights into stage-to-stage transitions.

Blood specimens were not collected at a set time of day, which led to fluctuations in measured testosterone within participants due to the diurnal variation in testosterone levels. Additionally, at lower Tanner stages, testosterone was often below detectable limits. Thus, raw testosterone levels could not be reliably used as an objective measure of sexual maturity. Instead, we modeled log-transformed testosterone values as a function of age using a simple mixed-effects model, incorporating a second-degree polynomial for age to account for potential non-linear trends. Individual-level smoothed predicted testosterone values were then generated from this model.

Generalized estimating equations (GEE) were used to model the probability of achieving maturity for each Tanner metric and axillary hair as a function of age, employing an exchangeable correlation structure to account for repeated measures within participants. Each Tanner metric was dichotomized for this analysis, assigning a value of 1 to the highest possible stage and 0 to all lower stages.

To assess the predictive value of individual Tanner metrics, ordinal logistic regression modeled overall Tanner staging (Stages I-IV) using single and combined metrics. Additional variables, including axillary hair, age, and testosterone levels, were included as predictors. Model performance was evaluated using the Area Under the Curve (AUC), measuring the discriminative power of each predictor, with values closer to 1 indicating stronger discriminative accuracy. All analyses were conducted using R Software version 4.3.2 and Stata Software version 15.1 (StataCorp, 2017).

## Results

### Participant Demographics and Baseline Characteristics

A total of 200 sexually naive, uncircumcised adolescent boys were enrolled in the study, with a median baseline age of 16 years (IQR = 1). At enrollment, 77.5% had primary level education, and 62.5% were currently attending school. At baseline, 6.0% of participants were at Tanner stage II, 13.5% at stage III, 55.0% at stage IV, and 25.5% at stage V (Table 1). Additionally, 91.0% of participants remained in the study through year 3.

**Table 1:** Baseline Characteristics of the Study Population.

|  | N (%) |
| --- | --- |
| **Total** | 200 (100.0) |
| **Age, years** |  |
| 15 | 83 (41.5) |
| 16 | 80 (40.0) |
| 17 | 37 (18.5) |
| **Education** |  |
| None | 1 (0.5) |
| Primary school | 155 (77.5) |
| Secondary school | 44 (22.0) |
| **Primary occupation** |  |
| Student | 125 (62.5) |
| Agriculture | 34 (17.0) |
| Mechanic | 10 (5.0) |
| Other* | 31 (15.5) |
| **Tanner stage** |  |
| I | 0 (0.0) |
| II | 12 (6.0) |
| III | 27 (13.5) |
| IV | 110 (55.0) |
| V | 51 (25.5) |
| **Follow up visits, median (range)** | 11 (0,12) |

\* Other: Construction, fishing, carpentry, metalwork

Axillary hair assessments were incorporated into the study and as such, a total of 517 (24.7%) data points were not collected, with participants missing a median of 3 assessments (IQR = 1; range: 1-4).

### Tanner Staging by Age and Single Metric Comparisons

The median ages at Tanner stages II, III, IV, and V were 16.2 years (IQR = 1.3), 16.3 years (IQR = 1.4), 17.4 years (IQR = 1.6), and 18.0 years (IQR = 1.8, respectively. We assessed the overall Tanner score by age and found a significant increase with age (Kendall’s τ = 0.292, p < 0.001) (Fig 1). Next, we examined the association between individual Tanner metrics and age, identifying pubic hair as having the strongest correlation (Kendall’s τ = 0.423, p < 0.001), while testicular volume showed the weakest (Kendall’s τ = 0.158, p < 0.001). Interestingly, axillary hair, despite not being a Tanner metric, exhibited a strong positive association with age (Kendall’s τ = 0.441, p < 0.001). Testosterone levels also showed a significant positive correlation with age (Kendall’s τ = 0.433, p < 0.001).

**Fig 1:**
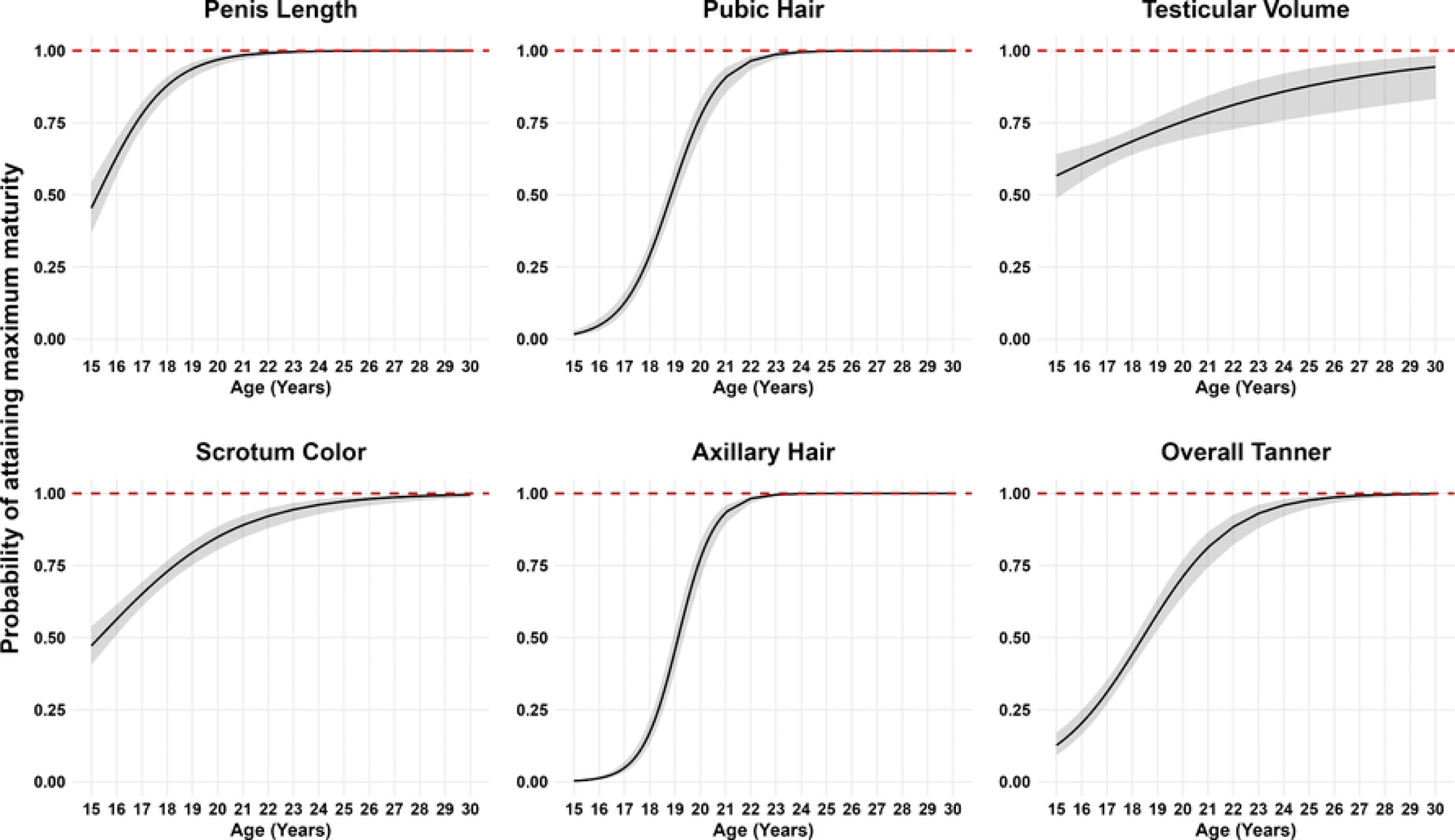
Tanner and Wolfsdorf Staging (axillary hair) by Age. Axillary hair assessments were introduced nine months into the study; therefore, data are missing for prior visits. Missing values for axillary hair were imputed using a linear mixed-effects model, incorporating age as a fixed effect and participant ID as a random intercept to account for individual variability in pubertal timing. The model was trained on participants with complete data and used to predict missing values. To facilitate comparison, both Tanner and axillary hair metrics were rescaled to a common 1-5 range.

### Probability of attaining maximum pubertal maturity

We modeled the probability of attaining maximum pubertal maturity for the Tanner metrics and axillary hair as a function of age using general estimating equations while accounting for clustering by participants. Fig 2 illustrates the predicted probability curves along with 95% confidence intervals for each trait across the age range of 15 to 30 years.

**Fig 2:**
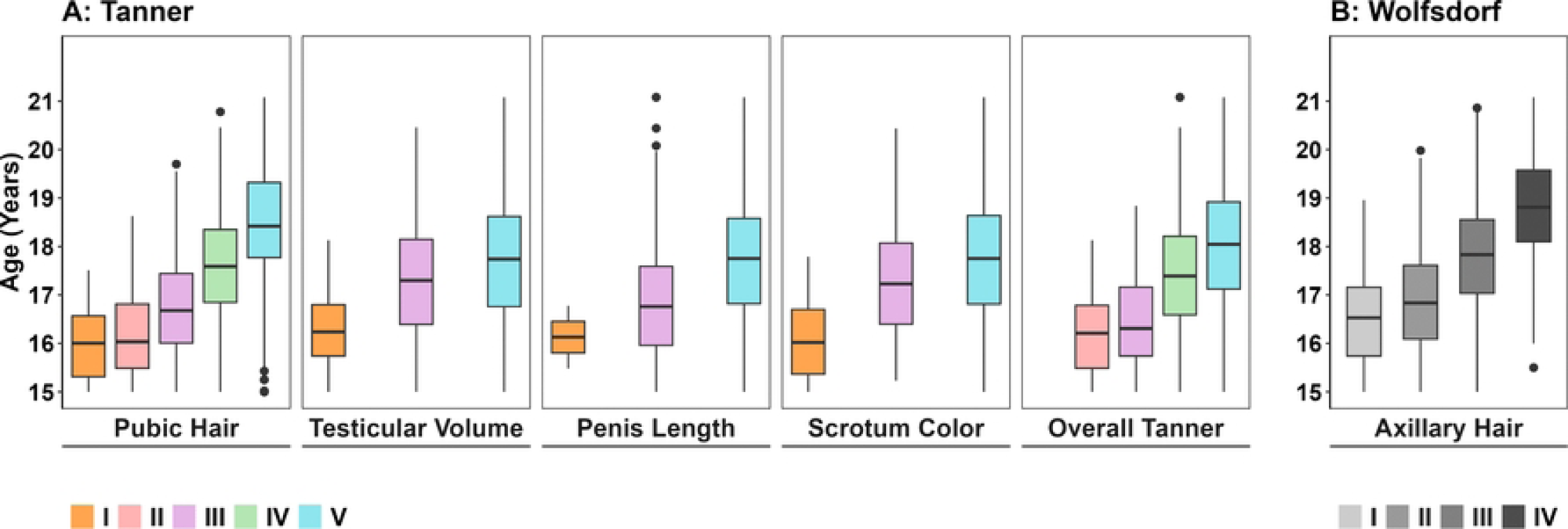
Predicted probability of attaining maximum pubertal maturity by age for each Tanner metric, overall Tanner score and axillary hair. Each panel shows predicted probabilities (black line) and 95% CIs (shaded area) from generalized estimating equation models accounting for clustering by participant for the probability of attaining maximum maturity across age (15–30 years) for each pubertal trait. A blue dotted line indicates full maturity (probability = 1.0).

The trajectory of pubertal maturation varied across the pubertal traits. Penis length, pubic hair, and axillary hair growth showed relatively early maturation trajectories, with steep increases in the probability of maximum maturity occurring between ages 15 and 20. These pubertal traits reached near-complete maturation by approximately age 21, with an over 95% chance of attaining maturity.

In contrast, testicular volume and scrotum color demonstrated more gradual maturation curves, with slower increases in the probability of full maturity. Near-maximum maturity for scrotum color was not reached until around age 25–27, while for testicular volume, maximum maturity was not reached by age 30.

The overall Tanner score, representing composite pubertal development, followed a curve slightly similar to that of the earlier-maturing traits, with a less steep transition between ages 15 and 22.

### Assessing Transitions Between Tanner Stages

Given the subjective nature of Tanner metric scoring and observed reversions to lower scores at some visits, we analyzed the dynamics of Tanner stage transitions using Markov transition probabilities. As expected, overall Tanner scores generally increased over the 3-monthly visits (Fig 3). The highest probability of forward transition (0.45) was observed between Tanner stages III and IV, while the lowest (0.25) occurred between stages II and III. Stages IV and V had the highest combined transition probability (0.59), indicating frequent back-and-forth movement between these stages. Individuals at stage V had the highest probability (0.69) of remaining at that stage, whereas stage III showed the lowest retention probability (0.41).

**Fig 3:**
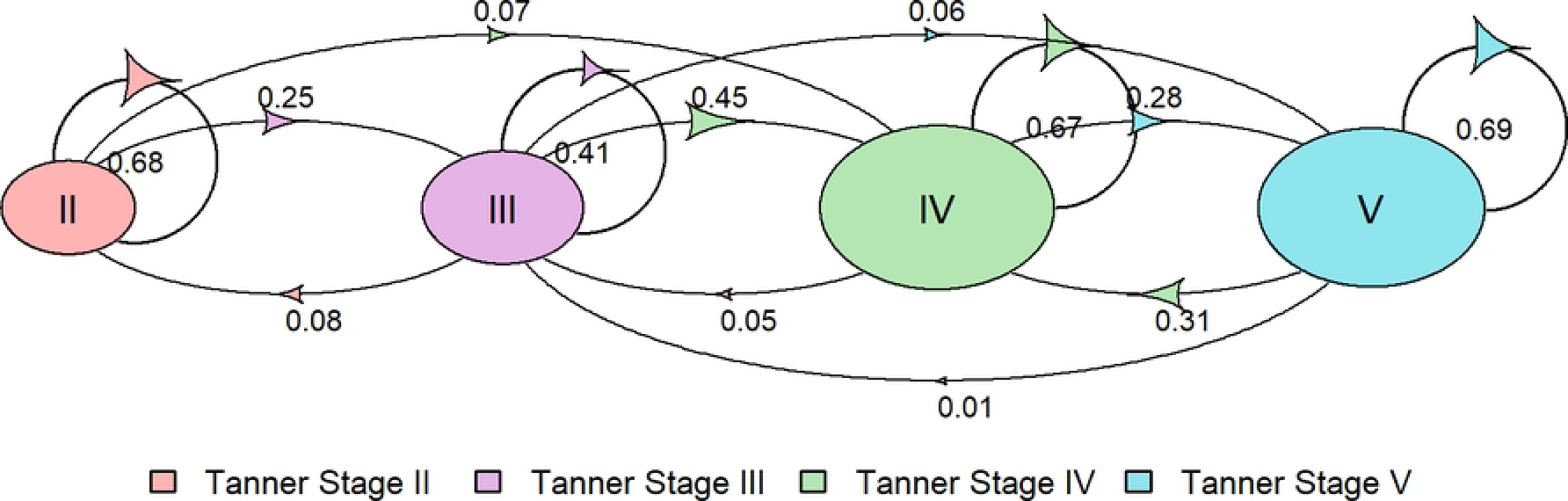
Markov transition probabilities between Tanner stages. Figure illustrates estimated probabilities of transitioning between Tanner stages over a single time interval, as modeled by a discrete-time Markov process. Each node represents a Tanner stage (Stage II to IV), and arrows between nodes indicate the direction and probability of transitioning from one stage to another. Arrow size corresponds to the magnitude of the transition probabilities, with larger arrows representing more likely transitions. Self-loops indicate the probability of remaining in the same stage.

### Prediction of Overall Tanner Score by Individual Metrics and Metric Combinations

We conducted an area under the curve (AUC) analysis to determine which single Tanner metric best predicted the overall Tanner stage. Pubic hair was the strongest predictor (AUC = 0.867, 95% CI [0.848, 0.885]), followed by testicular volume (AUC = 0.860, 95% CI [0.846, 0.876]) (Fig 4). Penis length had the lowest predictive value (AUC = 0.838, 95% CI [0.821, 0.853]), closely followed by scrotum color (AUC = 0.849, 95% CI [0.832, 0.866]). Among non-Tanner metrics, testosterone was the best predictor (AUC = 0.836, 95% CI [0.819, 0.857]), while age and axillary hair had AUC scores of 0.814 (95% CI [0.791, 0.834]) and 0.801 (95% CI [0.777, 0.823]), respectively.

**Fig 4:**
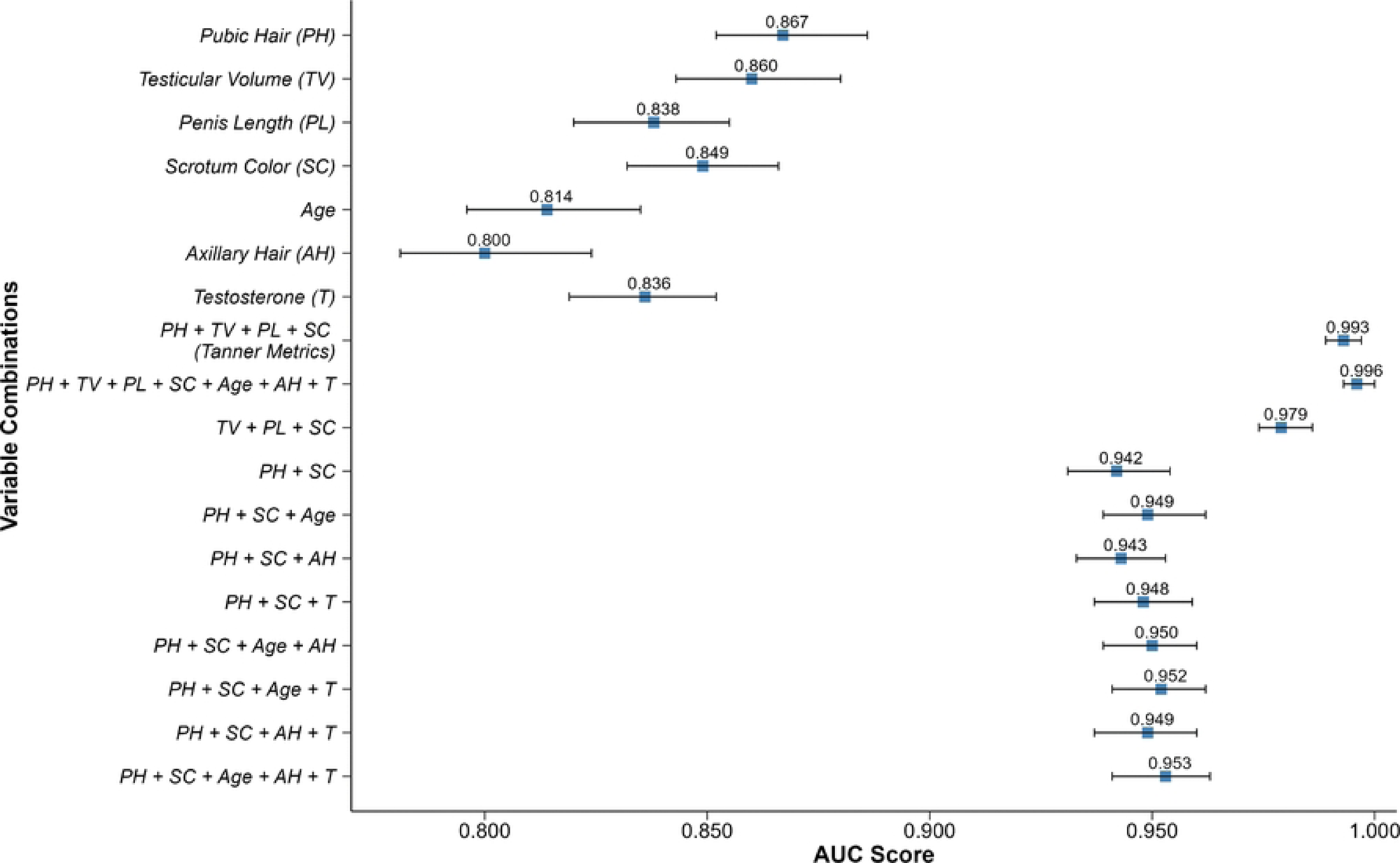
Area Under the ROC Curve values with 95% confidence intervals for models estimating discriminative accuracy for Tanner Stages using Tanner and non-Tanner metric combinations. This figure compares the discriminative performance of models incorporating both Tanner and non-Tanner metrics for Tanner stage classification. The central point on each bar indicates the estimated AUC, while the horizontal line shows the 95% confidence interval around that estimate. Higher AUC values indicate better model discrimination.

Evaluating combinations of Tanner metrics for overall Tanner stage prediction revealed, as expected, that the four-metric combination of pubic hair, testicular volume, penis length, and scrotum color had the highest AUC (0.993, 95% CI [0.990, 0.998]) (Fig 4). Including all Tanner and non-Tanner metrics only slightly improved the AUC to 0.996 (95% CI [0.993, 0.999]). Among three-metric combinations, testicular volume, penis length, and scrotum color yielded the highest AUC (0.979, 95% CI [0.974, 0.985]), followed by pubic hair, testicular volume, and scrotum color (AUC = 0.976, 95% CI [0.970, 0.983]). The lowest AUC among Tanner metric combinations was observed for pubic hair and penile length (0.920, 95% CI [0.906, 0.931]). However, the less intrusive pairing (i.e., not requiring physical manipulation of the genitals) of pubic hair and scrotum color yielded the highest AUC (0.942, 95% CI [0.930, 0.954]) among two-metric combinations. When considering the addition of non-Tanner metrics to the less intrusive characteristics, the combination of pubic hair, scrotum color, age and axillary hair resulted in 0.953 (95% CI [0.940, 0.960]). Adding serum testosterone marginally improved the AUC to 0.953 (95% CI [0.944, 0.963]).

### Prediction of Tanner Stage by Serum Testosterone Levels

Given the central role of testosterone in male puberty, we evaluated the predictive probabilities of each Tanner stage based on predicted serum testosterone levels. Serum testosterone levels below 50 ng/dL were most predictive of Tanner stage II (Fig 5). Between 50 and 250 ng/dL, although predicted probabilities remained relatively low (0.0 to 0.5), the stages were still distinguishable. Stage IV had the highest likelihood within this range, followed by Stage V. Meanwhile, probabilities for Stages II and III declined sharply, reflecting progression beyond the early stages. At testosterone levels above 250 ng/dL, the likelihood of predicting Stages IV and V increased markedly, suggesting a transition toward later pubertal development.

**Fig 5:**
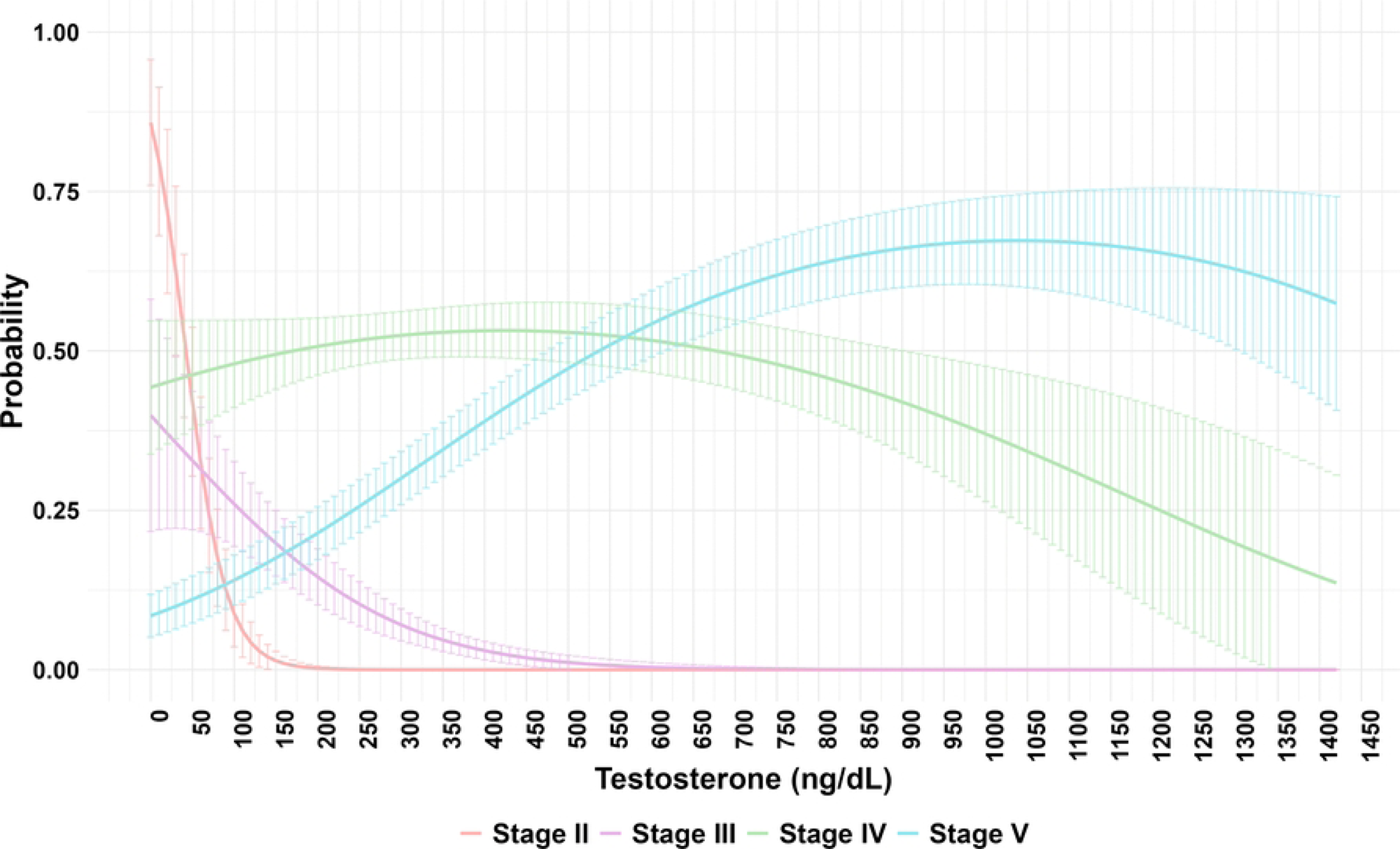
Predicted probabilities for Tanner Stages by Testosterone. The figure illustrates the model-estimated probabilities of being in each Tanner stage as a function of predicted testosterone concentration (ng/dL). Each curve represents the predicted probability for a specific Tanner stage, showing how the likelihood of being in that stage changes with increasing testosterone levels. Probabilities were derived from logistic regression, using robust standard errors clustered by individual.

## Discussion

Accurate determination of puberty stages is essential for assessing sexual maturity development and optimizing pediatric care, including addressing emerging behavioral concerns, mental health, dietary preferences among adolescents, non-communicable diseases and obesity that are on the rise worldwide [15–17]. The original Tanner staging system was developed in a European context, raising questions about the potential impact of regional or racial differences on Tanner staging performance and reliability in other populations, such as skin color variations affecting scrotal metric assessments. Additionally, classical Tanner staging involves observation and intrusive genital palpation during physical exams.

In this study, we assessed the performance of Tanner staging among African adolescent males and identified a set of minimally intrusive metrics as potential alternatives to the classic Tanner staging system. By combining key maturity parameters, such as pubic hair, scrotal color, age, and axillary hair, we found that these metrics can substantially reduce the intrusiveness of the clinical assessment while still maintaining 95% predictive accuracy.

As expected, we found that Tanner stage generally increased with age, consistent with the original published sources [6]. The probabilities of forward transition varied between Tanner stages, and we also observed several instances of reverse transitions. These variations may partly reflect examiner subjectivity in parameter scoring (e.g., changes in hair distribution between visits, especially during longitudinal assessments), underscoring the subjective nature of Tanner staging and need for examiner consistency. Including photographic capture, as was done in the original description [6], might help improve this consistency, although it would clearly add to the intrusiveness of the exam.

Considering individual Tanner metrics, pubic hair distribution showed the best discriminative ability for the overall Tanner score but misclassified approximately 13% of adolescents when used alone. Other Tanner metrics moderately predicted the overall Tanner score. Non-Tanner metrics, such as serum testosterone levels, age, and axillary hair, showed limited utility when used alone, performing worse than Tanner metrics.

Among combinations of Tanner metrics, the combination of testicular volume, penis length, and scrotum color best predicted the overall Tanner stage. A two-metric combination of pubic hair and scrotal color yielded a good predictive accuracy (marginally improved with the inclusion of axillary hair and age). This combination of pubic hair and scrotum color performed well as a minimally intrusive approach to maturity scoring. Notably, scrotal color was predictive even in this population with dark skin, prior to the onset of puberty.

Despite being a key driver of puberty onset and male maturity, serum testosterone levels had low overall predictive probabilities for all Tanner stages in our study population. Nevertheless, low serum testosterone levels were most predictive of Tanner stage II compared to stages III to V. We were unable to assess any associations with Tanner stage I, as most participants were past this stage at enrollment. Although testosterone alone may not reliably distinguish among adjacent Tanner stages, it appears useful in differentiating early from late pubertal development, with the most rapid transition occurring within intermediate testosterone levels (e.g., 75ng/dL to 250ng/dL).

Taken together, these findings indicate that pubertal staging approaches that substantially reduce or eliminate genital manipulation can retain high discriminatory accuracy for pubertal maturation. In longitudinal research, population-based surveillance, and clinical contexts where genital examination is impractical, poorly accepted, or ethically challenging, a minimally intrusive assessment combining pubic hair distribution, scrotal color, axillary hair, and age may reasonably replace traditional Tanner staging without meaningful loss of information.

## Limitations

This was a retrospective analysis of data collected to assess the effect of sexual debut on the penile microenvironment. As a result, the participant enrollment approach targeted males nearing the age of sexual debut and resulted in approximately 80% of participants having a baseline Tanner stage between IV and V, which may have limited our ability to robustly assess trends among lower Tanner stages. Continuous Tanner variables (e.g., penile length and testicular volume) were recorded as categorical rather than measured numerical values, potentially limiting more granular analysis of the associations. Additionally, the time-of-day blood samples were collected for serum testosterone testing was not standardized and likely resulted in highly variable serum testosterone levels within individual participants because serum testosterone levels vary throughout the day, being highest before 10 AM [18]. Standardizing collection times would have added substantial complexity, e.g. interruption of school hours, to study logistics and compliance. To address this, we used smoothed predicted testosterone values generated by modeling log-transformed testosterone values against age using a simple mixed-effects model.

## Conclusions

This study provides longitudinal evidence that pubertal maturation in East African adolescent males follows age-related trajectories similar to those originally described in populations of European descent [6], while also highlighting important limitations of traditional Tanner staging in real-world research settings. We identified an alternative combination of specific Tanner and non-Tanner metrics that provide a less intrusive sexual maturity scoring algorithm that has a predictive ability comparable to the original Tanner metrics. Future studies in younger adolescents, females, and diverse populations will be essential to determine the generalizability of these findings and to support broader adoption of context-appropriate pubertal staging methods.

## Data Availability

The data underlying this study cannot be made publicly available because they contain potentially identifiable participant information and are subject to the data-sharing policies of the Rakai Health Sciences Program (RHSP). De-identified data may be made available to interested parties upon reasonable request, subject to completion of the RHSP Data Request Form and execution of a Data Transfer Agreement. Requests for data access should be directed to Anthony Ndyanabo at

## Acknowledgements

The authors wish to thank all study participants and their parents/guardians for consenting to study participation, as well as the entire study staff for their dedication to its implementation.

